# Sitagliptin decreases visceral fat and blood glucoses in women with polycystic ovarian syndrome

**DOI:** 10.1101/19001685

**Authors:** Jessica K. Devin, Hui Nian, Jorge E. Celedonio, Patricia Wright, Nancy J. Brown

## Abstract

**Context:** Women with polycystic ovarian syndrome (PCOS) have decreased growth hormone (GH), which can increase visceral adiposity (VAT) and impair vascular function. GH releasing hormone, a dipeptidyl peptidase-4 (DPP4) substrate, stimulates GH secretion.

**Objective:** We tested the hypothesis that DPP4 inhibition increases GH and improves glucose levels and vascular function in women with PCOS.

**Methods:** Eighteen women with PCOS participated in a double-blinded, cross-over study. They received sitagliptin 100 mg vs. placebo daily for one month separated by an eight-week washout. During each treatment, women underwent a 75-gram oral glucose tolerance test (OGTT), assessment of vascular function and body composition. Overnight GH secretion was assessed via venous sampling every 10 minutes for 12 hours and analyzed using an automated deconvolution algorithm.

**Results:** During OGTT, sitagliptin increased GLP-1 (p<0.001), early insulin secretion (from mean insulinogenic index 1.9±1.2 (SD) to 3.2±3.1; p=0.02) and decreased peak glucose (mean −17.2 mg/dL [95% CI −27.7, −6.6]; p<0.01). At one month, sitagliptin decreased VAT (from 1141.9±700.7 to 1055.1±710.1 g; p=0.02) but did not affect vascular function. Sitagliptin increased GH half-life (from 13.9±3.6 to 17.0±6.8 min, N=16; p=0.04) and interpulse interval (from 53.2±20.0 to 77.3±38.2 min, N=16; p<0.05) but did not increase mean overnight GH (p=0.92 vs. placebo).

**Conclusions:** Sitagliptin decreased the maximal glucose response to OGTT and VAT. Sitagliptin did not increase overnight GH but increased GH half-life and the interpulse interval.

**Precis:** Sitagliptin improved body composition and blood glucoses following oral glucose load in women with PCOS. Sitagliptin potentiated GH half-life but did not increase overnight GH levels.

## Introduction

Growth hormone (GH) is secreted in a pulsatile fashion from the pituitary gland and exerts its effects either directly, via activation of the GH receptor, or indirectly, through hepatic insulin-like growth factor-1 (IGF-1). In adults, GH has important effects on the vasculature and body composition. GH increases endothelium-dependent vasodilator function and reduces inflammation.(1-3) We have previously shown that patients with low GH have impaired conduit artery vasodilator function along with decreased tissue-type plasminogen activator (tPA) activity and a defective fibrinolytic response to venous occlusion.(4) Others have corroborated these findings and further demonstrated that these vascular indices improve with GH replacement therapy.(3,5) GH also has anabolic and lipolytic effects. Conversely, GH secretion is reduced in obesity and particularly in individuals with visceral adiposity. Women with increased visceral adipose tissue (VAT) demonstrate a four-fold reduction in mean GH levels as compared to those without.(6) Normalization of GH secretion in patients with diminished GH affords one potential mechanism to address simultaneously both VAT and cardiovascular risk. Unfortunately, therapy with recombinant GH does not restore pulsatile secretion, is not subject to physiologic negative feedback by IGF-1, and is limited by hyperglycemia.(7)

We propose that an alternative strategy to enhance endogenous GH secretion in humans is to inhibit the degradation of endogenous GH releasing hormone (GHRH) by the dipeptidyl peptidase-4 (DPP4) enzyme. GHRH is the primary stimulus for pituitary GH secretion and determines GH pulsatility. Endogenous GHRH has a half-life of six minutes as it is degraded and inactivated by DPP4.(8,9) Sitagliptin was the first oral DPP4 inhibitor approved by the US Food and Drug Administration for the management of hyperglycemia in patients with type 2 diabetes mellitus. Sitagliptin improves post-prandial hyperglycemia in a glucose-dependent manner by decreasing the degradation of the incretin hormone, glucagon-like peptide-1 (GLP-1), and can therefore be safely given to patients without diabetes mellitus.(10) We recently found that acute DPP4 inhibition with sitagliptin enhances stimulated GH secretion and free IGF-1 levels in healthy, lean women. Moreover, while vasodilation and tPA activity levels increased in both men and women during stimulated GH secretion, women demonstrated an enhanced vascular response to increased GH.(11) The effect of chronic DPP4 inhibition on pulsatile GH secretion in individuals with low GH levels has not been studied.

Women with polycystic ovarian syndrome (PCOS) have decreased GH secretion characterized by an impaired GH response to stimuli and a greater than 50% reduction in 24-hour mean GH levels.(12-14) Approximately ten percent of reproductive-aged women in the United States are affected by PCOS at an estimated annual cost of over four billion dollars.(15) These women are often overweight and are at increased risk for future cardiovascular disease and diabetes mellitus. There are currently no approved medical therapies for women with PCOS; management options include weight loss or medications aimed at controlling symptoms of hirsutism or oligomenorrhea, or treatment of insulin resistance.(16,17) In this study, we tested the hypothesis that one month of DPP4 inhibition with sitagliptin would enhance overnight pulsatile GH secretion in overweight women with PCOS and improve glucose metabolism. Given the effects of GH on the vasculature, we also hypothesized that an increase in GH would enhance endothelial function and fibrinolysis.

## Materials and Methods

### Study Protocol

Eighteen women (BMI ≥25 kg/m^2^) with PCOS, 18 through 45 years of age completed a double-blind, randomized, placebo-controlled, crossover study. (See **Table 1** for subject characteristics and **Figure 1** for participant flow diagram.) The study adhered to the principles of the Declaration of Helsinki and Title 45, U.S. Code of Federal Regulations, Part 46, Protection of Human Subjects and was approved by the Vanderbilt University Medical Center Institutional Review Board. All subjects provided written informed consent prior to initiation of study procedures. A diagnosis of PCOS was confirmed when women met two out of the three 2003 Rotterdam criteria: oligomenorrhea (menstrual cycles occurring at intervals >35 days, or only 4-9 cycles per year) or secondary amenorrhea (no cycle in 3 months if previously regular or no cycle in 9 months if previous oligomenorrheic); clinical and/or laboratory evidence of hyperandrogenism; previous evidence of polycystic ovaries on ultrasound examination. Other common causes of hyperandrogenemia and oligomenorrhea, including hyperprolactinemia, late-onset 21-hyroxylase deficiency, and inadequately treated hypothyroidism were excluded at the time of screening visit.(18) Patients with a history of chronic illness, including diabetes mellitus, hypertension, cardiovascular disease, and chronic renal or hepatic insufficiency as well as a history of weight loss surgery or ongoing night-shift work were excluded. Women were required to discontinue oral contraceptives for eight weeks and spironolactone for thirty days prior to study enrollment, if applicable. One woman took a stable dose of metformin throughout the study; other drugs known to alter glucose or insulin metabolism were not permitted. Pregnancy was excluded in all women of child-bearing age by serum pregnancy testing prior to study drug initiation and again prior to performing dual-energy x-ray absorptiometry (DEXA).

**Table 1.**
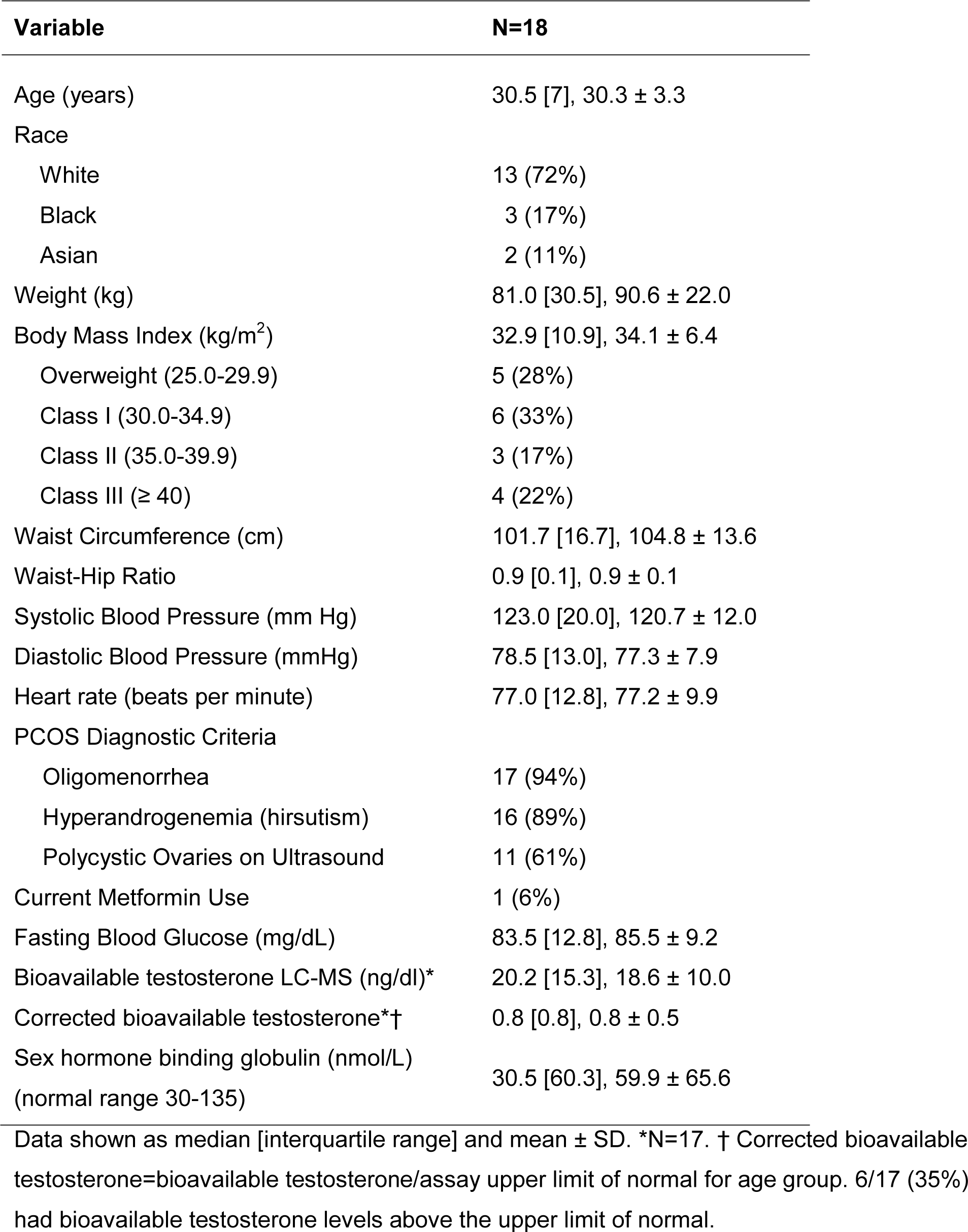
Subject Characteristics.

**Figure 1.**
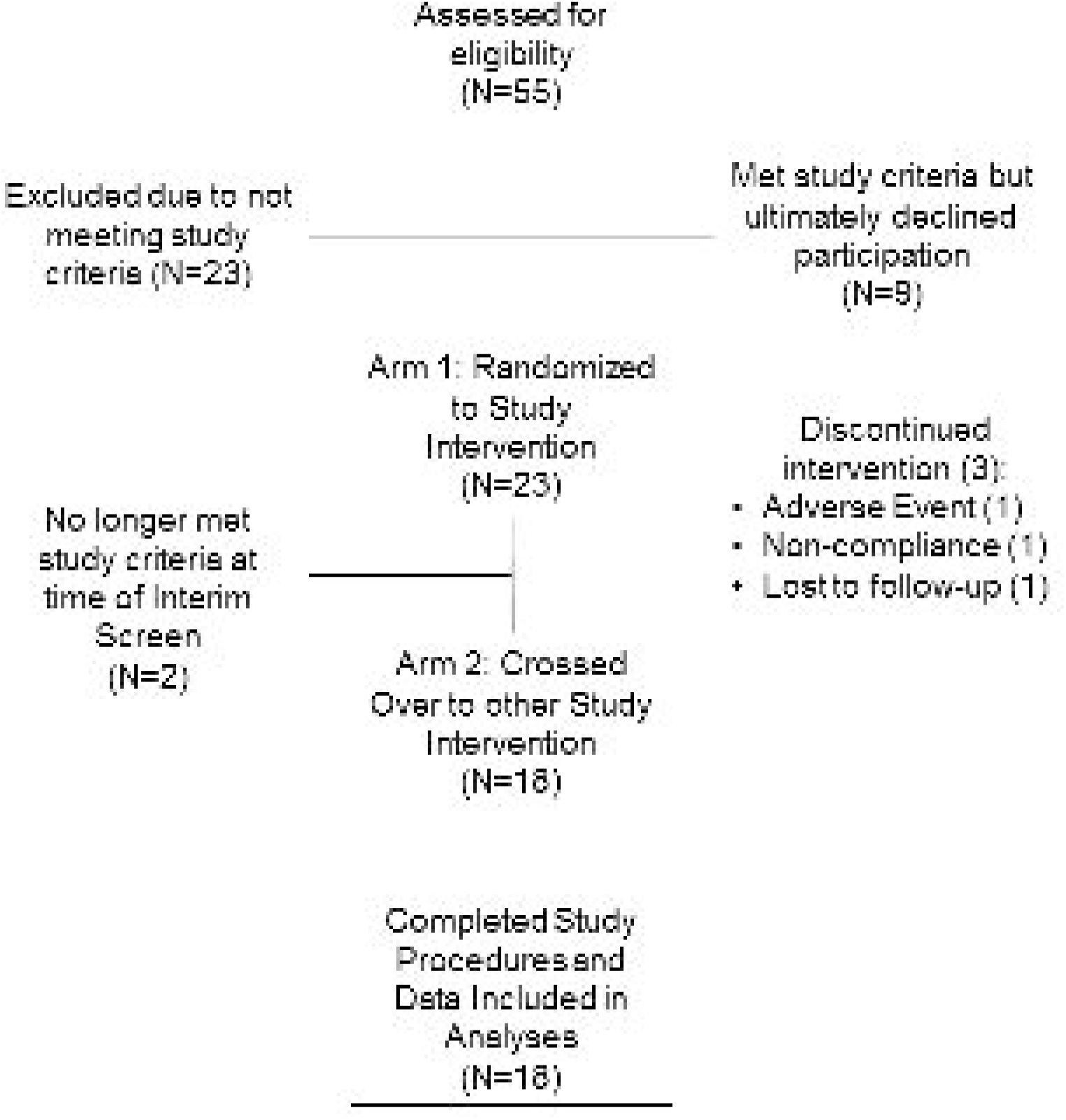
Participant Flow Diagram. Fifty-five women with polycystic ovarian syndrome (PCOS) were screened for study participation and 32 were determined to be eligible. Twenty-three women agreed to participate and were randomized to study drug. Eighteen of these women participated in both study arms and their data were included in analyses. Complete data was available only from both outpatient visits in one woman. Another woman did not complete the overnight GH sampling portion of both inpatient visits.

Women underwent two one-month treatment periods separated by wash-out period of at least eight weeks (**Figure 2**). Subjects were assigned to treatment order (double-blinded sitagliptin 100 mg p.o. qd or matching placebo) using a block randomization algorithm. On each outpatient and inpatient study day, subjects reported to the Vanderbilt Clinical Research Center (CRC) in the morning after an overnight fast. After approximately two weeks of study drug, subjects underwent an extended OGTT. One hour after oral study drug (sitagliptin vs. placebo) a peripheral intravenous line was placed in the antecubital fossa of the non-dominant arm. Baseline venous samples and vital signs were obtained at least 60 minutes following study drug. Oral glucola (75 grams) was then ingested within 10 minutes, and samples were obtained over the following 270 minutes. Women were then discharged and continued daily study drug at home for an additional two weeks.

**Figure 2.**
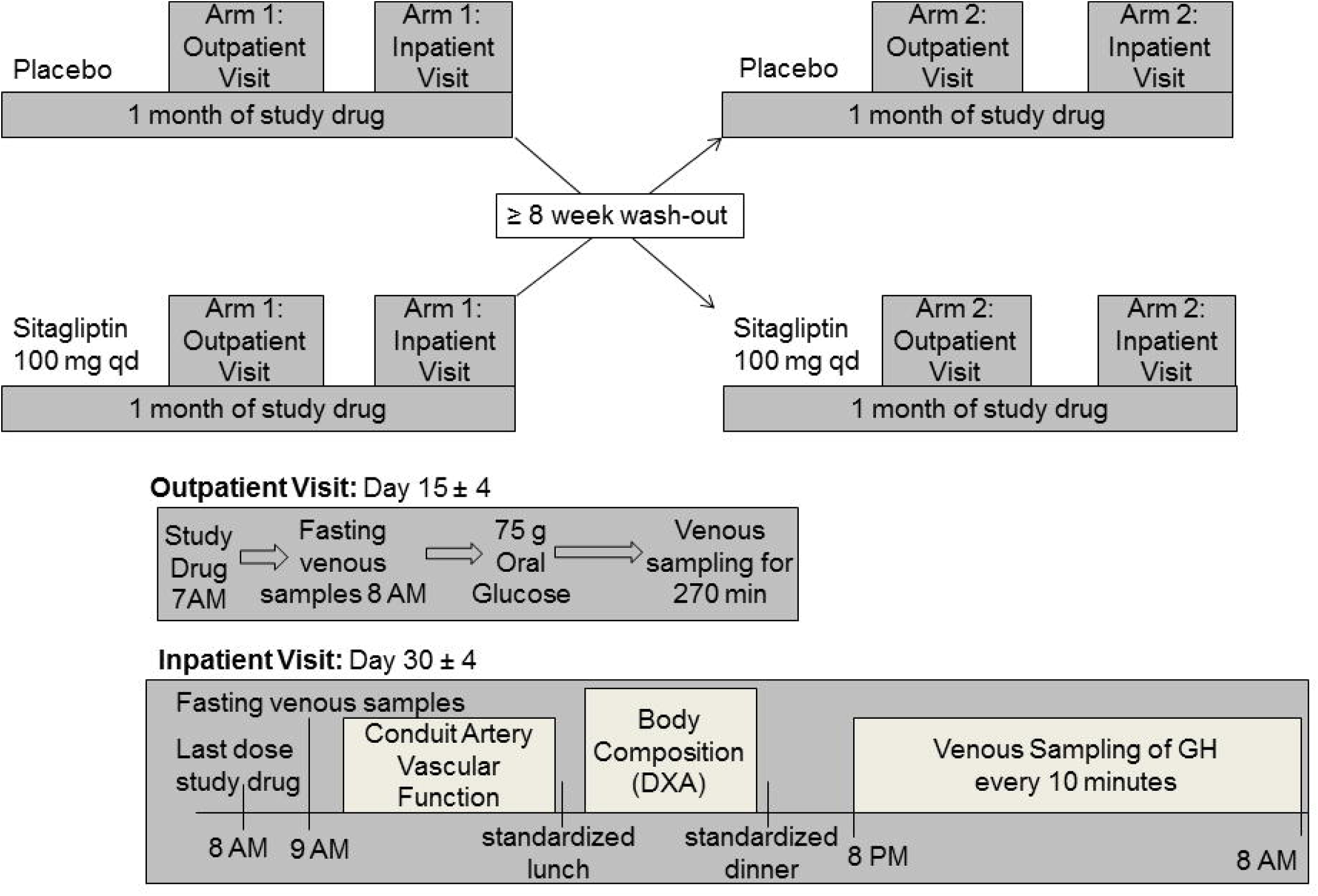
Subjects participated in a double-blind, randomized, placebo-controlled, crossover study. Eighteen women underwent two one-month treatment periods (sitagliptin 100 mg p.o. qd or matching placebo) separated by a washout period of at least eight weeks. On each study day, subjects reported to the Vanderbilt Clinical Research Center (CRC) in the morning after an overnight fast. After approximately two weeks of study drug, subjects underwent an extended OGTT (75-grams oral glucola). Women were then discharged and continued daily study drug at home for an additional two weeks. On the last day of study drug, women (N=17) again reported to the CRC for collection of fasting venous samples, measurement of endothelium-dependent and -independent vasodilation, measurement of total and regional body composition via duel-energy x-ray absorptiometry, and frequent venous sampling for GH every ten minutes for twelve hours overnight.

On the last day of study drug, women again reported to the CRC following an overnight fast. One hour after the last dose of oral study drug, vital signs, weight, and fasting venous samples were obtained via peripheral stick in the non-dominant arm. Women then underwent assessment of endothelium-dependent and independent vasodilation in the dominant arm (see “conduit artery vascular function” below). In the afternoon, total and regional body composition was determined by a certified densitometrist using dual-energy X-ray absorptiometry (Lunar iDXA; GE Health Care, Madison, WI) with enCore software (version 13.6). Women also completed the previously validated polycystic ovarian syndrome questionnaire (PCOSQ).(19-21) In the evening, women underwent placement of an indwelling venous catheter and venous blood was sampled for GH every ten minutes from 8 PM until 8 AM. Meal content and composition were standardized across all subjects, and intake of the standardized meals was matched within each woman across study arms. Snacking and exercise were not permitted in the CRC. Only water intake was permitted during GH sampling.

### Conduit Artery Vascular Function

Endothelium-dependent vasodilation was evaluated in a temperature-controlled room in the fasting state after 15 minutes of rest in the supine position. Participants were asked to refrain from exercise, alcohol, caffeine, and anti-inflammatory medications for 24 hours preceding each measurement. Studies could not be coordinated with the phase of the menstrual cycle due to the infrequency or absence of cycles in the majority of our volunteers. Vascular images were obtained by a single ultrasound-trained technician using an L12-3 broadband linear array transducer attached to a high-resolution ultrasound machine (EPIQ7C; Phillips, Bothwell, WA). A longitudinal image (parallel to the artery) was acquired just proximal to the antecubital fossa of the dominant arm with the transducer positioned to optimize images of the near and far wall interfaces. Depth and gain settings were optimized to identify the interface between the lumen and blood vessel wall. Anatomical landmarks were documented while positioning the probe, to facilitate repositioning during repeated studies. A simultaneous electrocardiogram signal was recorded throughout the imaging. To assess endothelium-dependent vasodilation, brachial artery diameter was measured under basal conditions and during reactive hyperemia. Reactive hyperemia was achieved by inflating a pneumatic cuff on the upper arm to 200 mmHg for five minutes. The cuff was deflated and images of the artery were obtained continuously across the cardiac cycle for 180 seconds after cuff release. Baseline velocity time integral (VTI) was determined prior to cuff occlusion and hyperemic VTI was determined for ten beats post-cuff release via pulse-wave spectral Doppler recordings. Following a 15-minute rest period, the brachial artery was imaged again to re-establish basal conditions. Endothelium-independent vasodilation was then determined by administering 0.4 mg nitroglycerin sublingually. The brachial artery was imaged three minutes later for two minutes. Nitroglycerin was only administered if systolic blood pressure was at least 110 mm Hg, pulse at least 60 bpm, and with the participant’s consent (N=3 participants).

Acquisition and analysis of the DICOM-stored images was performed using software (Brachial analyzer 5.0) designed for this purpose by Medical Imaging Applications LLC (Iowa City, Iowa). The vessel wall-lumen interface was determined by derivative based edge detection following identification of the region of interest by the blinded investigator (JKD). The selected region of interest was consistent across study days for the same subject. The maximum diameter post-hyperemia was obtained using previously described custom design software which automatically calculates the time to peak and peak dilation.(22) This software implements the method validated by Green et al. which uses a smoothing algorithm to correct for changes in the brachial artery diameter during the cardiac cycle.(23) The percent change in diameter, following reactive hyperemia and nitroglycerin, was then calculated as percent vasodilation=[(peak diameter-baseline diameter)/baseline diameter]*100. The reactive hyperemia-induced increase in VTI relative to baseline was calculated as a ratio=hyperemia VTI/baseline VTI.

### Laboratory Analyses

All samples were obtained after the first 3 mL of blood were discarded. Blood samples were collected on ice, centrifuged immediately and plasma stored at −80°C in pre-specified aliquots until time of assay. All samples obtained from each subject during sitagliptin and placebo treatment were simultaneously analyzed with internal controls. DPP4 activity was assayed by incubating 20 µl sample in 80 µl assay buffer (0.1 M Tris at a pH of 8.0; Bachem, Torrance, CA) for 30 minutes at 37°C with colorimetric substrate [2 mM L-glycyl-L-prolyl p-nitroanilide hydrochloride (Sigma Aldrich, St. Louis, MO)] for a total reaction volume of 200 µL, as previously described.(24) DPP4 antigen concentration was determined by enzyme-linked immunoassay (ELISA) (eBioscience; San Diego, CA). The enzyme activity was assessed by measuring the increase in specific absorbance at 405 nm at 0, 15, and 30 minutes and was expressed as nmol/mL/min. Percent DPP4 inhibition was determined by the equation: [1-(DPP4 activity during sitagliptin/DPP4 activity during placebo)]*100. GH levels were determined by two-site immunoassay (Beckman Access Ultrasensitive human GH assay, Beckman Coulter, Inc.) performed on the Dxl automated immunoassay system (Beckman Instruments; Chaska MN) and calibrated against NIBSC WHO IS 98/574 with an analytic measurement range of 0.002 to 35 ng/mL. The first five subjects were analyzed at Brigham Research Assay Core (Boston, MA); the intra-assay CV was 1.48-11.26% and the inter-assay CV was 1.96-14.4%. The remaining subjects were analyzed at the Mayo Immunochemical Core Lab (Rochester, MN); the intra-assay CV was 3.5% at 2.50 ng/mL and 3.2% at 14.8 ng/mL. The inter-assay CV’s were 4.3% at 3.03 ng/mL, 5% at 7.23 ng/mL, and 4.8% at 13.62 ng/mL. All GH samples from each subject were analyzed via batch in the same lab. Free IGF-1 was determined using a commercially available ELISA (R & D systems; Minneapolis, MN) with a minimum detectable range of 0.015 ng/mL, intra-assay CV 3.6-5% and inter-assay CV 10.0-11.1%. Total IGF-1 was analyzed by Luminex® assay (Millipore Sigma; Burlington, MA), calibrated against the NIBSC WHO IS 02/254, with an intra-assay CV <10% and intra-assay CV<15%. IGF binding protein-1 (IGFBP-1) and IGFBP-3 were analyzed by commercially available ELISAs (RayBiotech; Norcross GA) with a minimum detectable concentration of 5 pg/mL for IGF-BP1 and 80 pg/mL for IGF-BP3. Both assays report an intra-assay CV <10% and inter-assay CV <12%. tPA activity and plasminogen activator inhibitor-1 (PAI-1) antigen were measured in blood collected in acidified citrate anticoagulant (TriniLIZE™ Stabilyte tubes, Tcoag; Bray Co. Wicklow, Ireland). TPA activity was analyzed using an ELISA calibrated against NIBSC WHO IS 98/714 (Oxford Biomedical Research, Oxford MS). PAI-1 antigen was analyzed using the TintElize PAI-1 antigen assay (Tcoag; Co. Wicklow, Ireland). Samples for analysis of active GLP-1, total peptide YY (PYY) and PYY 3-36, and glucagon were collected in EDTA collection tubes prepared with aprotinin and DPP4 inhibitor (Millipore Sigma; Burlington, MA). Active GLP-1 was analyzed in duplicate using the MILLEPLEX® MAP Human Metabolic Hormone Magnetic Bead Panel (Millipore Sigma; Burlington, MA) with an intra-assay CV of <10% and inter-assay CV <15%. Total PYY and PYY 3-36, glucagon, and insulin were analyzed in duplicate by radioimmunoassay (Millipore Sigma; Burlington, MA) with intra-assay CVs of 3.7%, 2.3%, 1.6%, and 3.3%, respectively. PYY 1-36 was calculated by subtracting PYY 3-36 from Total PYY. Estradiol was also analyzed by radioimmunoassay (MP Biomedicals; Santa Ana, CA) with an intra-assay CV of 3.9%. Blood glucoses were determined by Accu-Chek® Inform II bedside glucometer (Roche Diagnostics; Indianapolis, IN); the same glucometer was used on each outpatient study day. High-sensitivity C-reactive protein (hsCRP) was determined by commercially available ELISA (Hycult®Biotech; Plymouth Meeting, PA). Bioavailable testosterone was calculated from testosterone measured via quantitative high-performance liquid chromatography-tandem mass spectrometry (ARUP® Laboratories; Salt Lake City, Utah). Sex hormone binding globulin was obtained via quantitative electrochemiluminescent immunoassay (ARUP® Laboratories; Salt Lake City, Utah). F2-isoprostanes were determined via gas chromatography/mass spectrometry assay, as previously described.(25) Total plasma cholesterol and triglycerides were measured by standard enzymatic assays. HDL cholesterol was measured via the enzymatic method after precipitation of VLDL and LDL using polyethylene glycol reagent. From these data LDL cholesterol was calculated using the Friedewald equation. Plasma free fatty acids were analyzed with a commercially available enzymatic kit (Wako Life Sciences; Richmond, VA). The insulinogenic index, quantitative insulin sensitivity check index (QUICKI), homeostatic model assessment of insulin resistance (HOMA-IR) and Matsuda Index were calculated as previously described. (26,27)

### Analysis of Growth Hormone Secretion

Overnight spontaneous GH secretion was assessed using an automated deconvolution algorithm (AutoDecon Pulse_XP Software Package), which determines secretion events, adds them to the current fit to test for statistical significance, and automatically removes any non-significant events until no additional statistically significant secretion events are found.(28) AutoDecon has been validated for the analysis of endogenous pulsatile GH secretion from time-series data obtained via frequent venous sampling from an indwelling catheter every ten minutes for twelve hours.(29,30) Initial algorithm estimates included the standard deviation of the secretion events, set at one-half of the data-sampling interval (5 min), and a starting value for the GH elimination half-life of 13 minutes based upon previously published data in adults with BMI >25 kg/m^2^.(31) Initial values for basal secretion and concentration at t=0 were also estimated from the secretion and concentration panels. AutoDecon then determined GH basal secretion, half-life, median pulse mass and interpulse interval, number of GH peaks, and GH area under the curve. Additional parameters of GH secretion including mean overnight GH secretion, GH peak and nadir were determined via Microsoft Excel 2010. Missing data were omitted from analyses.

### Statistical Analysis

Data are presented in results tables as both mean ± standard deviation and median [interquartile range, IQR], given that data is not normally distributed. We tested for carry-over effect using the T-test approach proposed by Jones and Kenward.(32) Repeated measurement continuous variable data from OGTT were summarized as mean over time (from baseline to 270 minutes) or area-under-the-curve (AUC) for the same time period. Unless otherwise noted, a paired t-test was used to compare variables between treatment conditions with results presented as the mean difference between treatments with 95% confidence interval (CI). Mixed effect models were also used to analyze repeated measures data during the OGTT with a random subject effect and with fixed effects of treatment (sitagliptin vs placebo), time and treatment x time interaction. The baseline measurement was also included in each model. Interaction terms were removed from the final model when the corresponding p value was >0.2. Comparisons between treatments were made at each time-point using Wilcoxon signed rank test. Spearman correlation was used to evaluate the association between continuous variables. Analyses were performed using IBM SPSS software v. 23.0, GraphPad Prism 5 and R 2.15.0 (www.r-project.org). A p-value less than or equal to 0.05 was determined to be significant.

### Sample Size Calculation

Sample size was calculated with Power and Sample Size Calculation (PS) Software using the design for a paired t-test.(33) In a prior study mean baseline overnight GH was 0.7 ± SEM 0.2 μg/L versus a mean overnight GH after therapy of 1.2 ± SEM 0.3 μg/L (p<0.005) in 13 subjects.(31) With the enrollment of subjects in current study, we found that the standard deviation (SD) of mean overnight GH was 0.51 µg/L (measurements from two periods combined), well below the SD of 0.72 µg/L and 1.1 µg/L observed in the prior study used in the original sample size calculation. Conservatively assuming a SD of 0.6 µg/L in each group, we calculated that a sample size of 16 women provides 93.2% power to detect a difference in GH means of 0.5 µg/L.

## Results

### Effect of sitagliptin on DPP4 activity and glucose metabolism

Seventeen women completed the entire study. One woman completed two weeks of both treatments and is also included in the analysis of the outpatient study days.(Figure 1) Sitagliptin 100 mg daily significantly decreased DPP4 activity (p<0.001 vs. placebo), as assessed on both the outpatient visit days (from 21.3 ± 8.0 to 8.8 ± 5.2 nmol/mL per minute) and the inpatient visit days (from 22.9 ± 6.0 to 9.9 ± 4.9 nmol/mL per minute).

Sitagliptin decreased blood glucose following ingestion of 75 grams of oral glucose, as summarized as both average over time (mean difference −5.13 mg/dL, 95% CI [-10.26, 0.00]; p=0.05), and AUC (mean difference −1476.25 mg/dL x 120 min, 95% CI [-2472.69, −479.81]; p<0.01) (**Figure 3A**). Peak blood glucose following the oral glucose load was also lower during sitagliptin treatment (mean difference −17.2 mg/dL, 95% CI [-27.7, −6.6]; p<0.01). The overall effect of sitagliptin on metabolism of glucose varied with time (p<0.001). At 100 minutes, mean blood glucose was 15.4 mg/dL (95% CI 8.7, 22.1; p<0.001) lower during sitagliptin treatment than during placebo.

**Figure 3.**
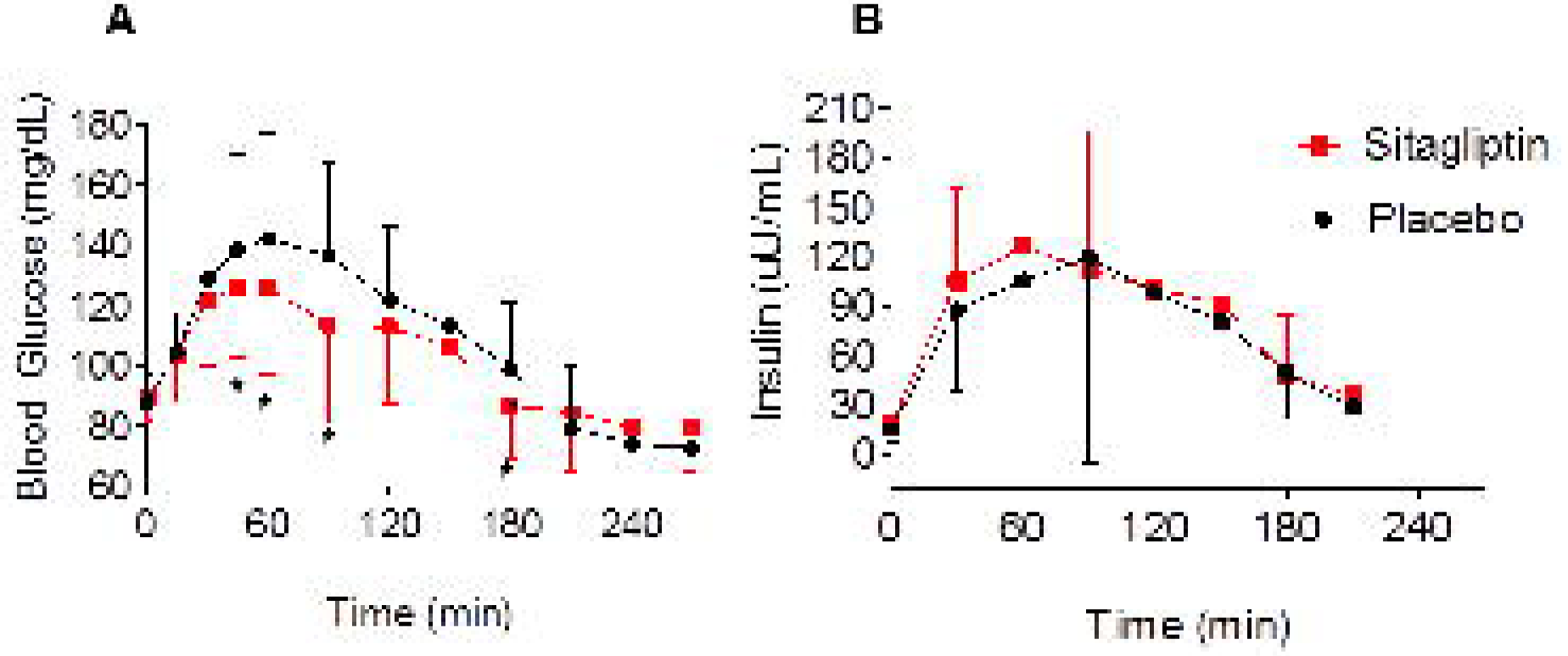
Sitagliptin improves blood glucoses (A) and early insulin secretion (B) following 75 grams oral glucose. Data were obtained on the outpatient visit day in 18 women and are presented as mean ± SD. The overall effect of sitagliptin on glucose levels varied with time (p<0.001). P value was obtained via mixed effect model with a random subject effect and with fixed effects of treatment (sitagliptin vs placebo), time and treatment x time interaction. *p ≤ 0.05 vs. placebo at corresponding timepoint, as obtained by Wilcoxon signed rank.

Sitagliptin also enhanced early insulin secretion, as demonstrated by an increase in the insulinogenic index (from 1.9 ± 1.2 to 3.2 ± 3.1; p=0.02) and insulin AUC for the first 30 minutes after glucose ingestion (mean difference 349.21 µU/mL x 30 min, 95% CI [39.3, 659.1]; p=0.05) (**Figure 3B**). Sitagliptin did not affect peripheral insulin resistance (Matsuda), hepatic insulin resistance (HOMA-IR) or insulin sensitivity (QUICKI), as determined on the day of the 75-gram OGTT (**Table 2**). Sitagliptin did not affect fasting blood glucose (p=0.572 vs. placebo) or fasting insulin levels (p=0.687 vs. placebo).

**Table 2.**
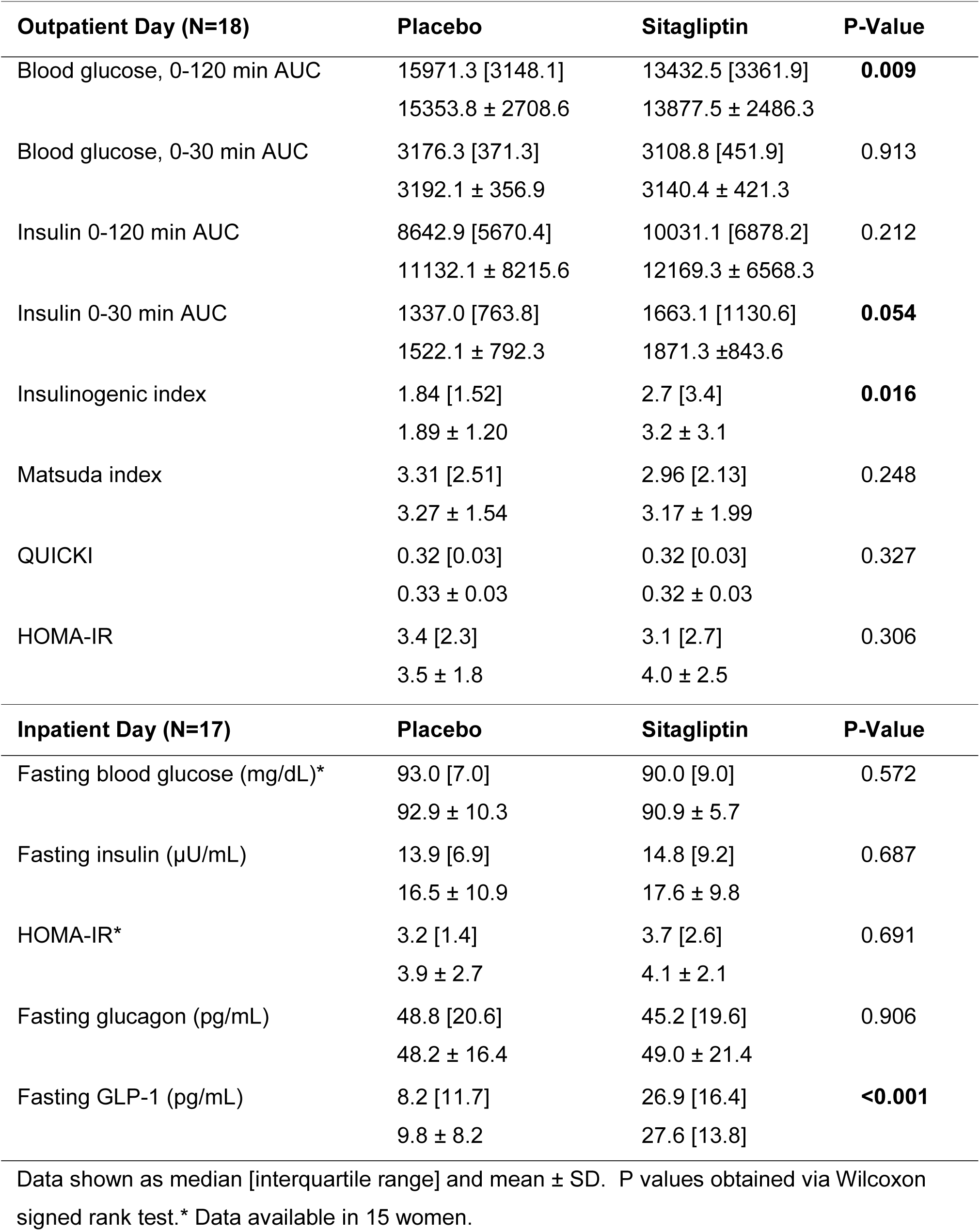
Effect of sitagliptin on glucose metabolism.

Sitagliptin increased fasting GLP-1 levels (p<0.001 vs. placebo) as well as GLP-1 levels during the 75-gram OGTT, summarized as average over time (mean difference 27.01 pg/mL, 95% CI [18.01, 36.01]; p<0.001) (**Figure 4A**). Sitagliptin also increased peptide YY 1-36 (mean difference 14.97 pg/mL, 95% CI [2.30, 27.63]; p=0.02) and decreased peptide YY 3-36 (mean difference −39.78 pg/mL, 95% CI [-51.39, −28.17]; p<0.001). (**Figure 4B and C**) Sitagliptin did not influence the change in free fatty acid levels or glucagon levels following oral glucose ingestion (data not shown).

**Figure 4.**
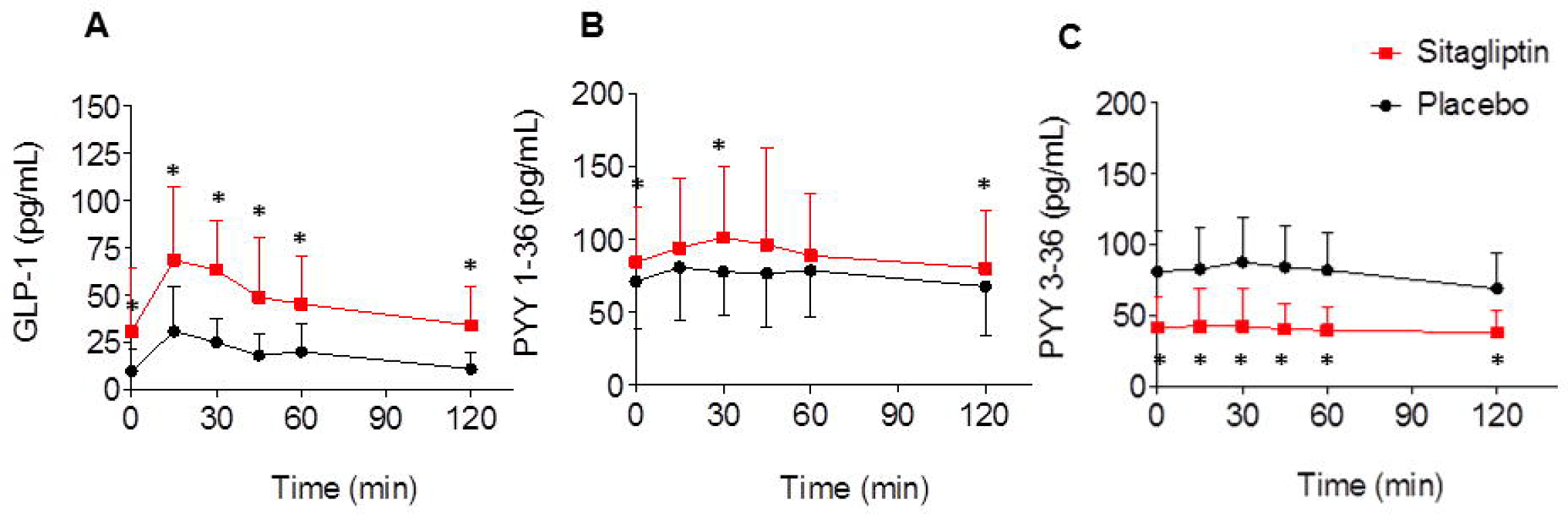
Sitagliptin increases both GLP-1 (A) and PYY 1-36 (B) and decreases PYY 3-36 (C) levels following oral 75 grams glucose. Data were obtained on the outpatient visit day in 18 women, and are expressed as mean ± SD. Model based p values for treatment effect are: p<0.001 for effect of sitagliptin on GLP-1, p<0.001 for effect of sitagliptin on PYY 1-36, p<0.001 for effect of sitagliptin on PYY 3-36. P value was obtained via mixed effect model with a random subject effect and with fixed effects of treatment (sitagliptin vs placebo), time and treatment x time interaction. *p ≤ 0.05 vs. placebo at corresponding timepoint, as obtained by Wilcoxon signed rank.

### Effect of sitagliptin on GH secretion and IGF-1 levels

Overnight GH secretion (GH AUC) correlated inversely with weight (placebo: r_s_= −0.69, p=0.003; sitagliptin: r_s_=-0.71, p=0.002), VAT mass (placebo: r_s_= −0.82, p<0.001; sitagliptin: r_s_= −0.77, p=0.001), high-sensitivity C-reactive protein (placebo: r_s_= −0.53, p=0.035; sitagliptin: r_s_= −0.54, p=0.031), and HOMA-IR (placebo: r_s_= −0.54, p=0.033; sitagliptin: r_s_=-0.79, p=0.001), regardless of drug treatment during the preceding month.

GH levels were appropriately suppressed following oral glucose ingestion and then increased approximately 4 hours later. Sitagliptin did not significantly influence the late GH peak (mean difference 0.31 ng/mL, 95% CI [-0.92, 1.54]; p=0.61) during the 75-gram OGTT. IGFBP-1 levels, summarized as average of time, were not affected by sitagliptin (mean difference −9.18 pg/mL, 95% CI [-104.73, 86.38]; p=0.84). Free IGF-1 levels also were unaffected by sitagliptin (mean difference −0.01 ng/mL, 95% CI [-0.09, 0.07]; p=0.84). Free IGF-1 levels at each time point following oral glucose were: baseline 0.46 ± 0.22 ng/mL placebo vs. 0.44 ± 0.21 ng/mL sitagliptin; 30 minutes after oral glucose 0.46 ± 0.19 ng/mL placebo vs. 0.43 ± 0.18 ng/mL sitagliptin; 90 minutes after oral glucose 0.44 ± 0.20 ng/mL placebo vs. 0.46 ± 0.20 ng/mL sitagliptin; 150 minutes after oral glucose 0.49 ± 0.26 ng/mL placebo vs. 0.45 ± 0.22 ng/mL sitagliptin.

During overnight sampling after one month of sitagliptin, GH half-life (p=0.04 vs. placebo) and consequently the median interpulse interval (p<0.05 vs. placebo) both increased, but GH AUC and mean GH were stable (**Table 3**). Representative plots of overnight sampling of GH from 3 subjects are included in **Figure 5**. One month of daily sitagliptin had no effect on fasting free IGF-1 levels, total IGF-1 levels, or IGFBP-3 levels (**Table 3**).

**Table 3.** Effect of sitagliptin on overnight GH secretion and IGF-1 levels.

| Variable (N=16) | Placebo | Sitagliptin | P-Value |
| --- | --- | --- | --- |
| GH AUC (ng/mL per 12 hr) | 428.9 [543.5]<br>604.5 ± 398.7 | 524.6 [466.0]<br>593.5 ± 371.6 | 0.918 |
| Basal GH secretion (ng/mL·min)* | 0.001 [0.002]<br>0.002 ± 0.002 | 0.002 [0.002]<br>0.002 ± 0.001 | 0.496 |
| GH half-life (min) | 13.9 [2.7]<br>13.9 ± 3.6 | 14.5 [8.0]<br>17.0 ± 6.8 | **0.044** |
| Number GH secretion events | 10.5 [4.0]<br>11.0 ± 2.6 | 8.5 [6.5]<br>9.1 ± 3.3 | 0.058 |
| Median secretion pulse mass | 1.1 [1.2]<br>2.2 ± 3.4 | 1.3 [1.8]<br>1.7 ± 1.5 | 0.756 |
| Median secretion pulse height | 0.04 [0.05]<br>0.10 ± 0.14 | 0.07 [0.16]<br>0.34 ± 0.63 | 0.301 |
| Median secretion interpulse interval | 51.1 [34.4]<br>53.2 ± 20.0 | 61.8 [60.6]<br>77.3 ± 38.2 | **0.049** |
| Maximum GH (ng/mL) | 4.1 [5.5]<br>5.1 ± 3.5 | 4.0 [3.5]<br>5.1 ± 4.4 | 0.918 |
| Minimum GH (ng/mL) | 0.03 [0.03]<br>0.04 ± 0.02 | 0.04 [0.05]<br>0.04 ± 0.02 | 0.567 |
| Mean GH (ng/mL) | 0.63 [0.73]<br>0.85 ± 0.54 | 0.74 [0.73]<br>0.84 ± 0.51 | 0.918 |
| Total IGF-1 (ng/mL)† | 81.4 [57.0]<br>88.7 ± 28.6 | 86.6 [52.1]<br>88.8 ± 29.7 | 0.586 |
| Free IGF-1 (ng/mL)† | 0.31 [0.27]<br>0.40 ± 0.26 | 0.38 [0.18]<br>0.43 ± 0.25 | 0.523 |
| IGFBP-1 (pg/mL) | 184.4 [149.4]<br>263.6 ± 298.3 | 158.2 [469.6]<br>325.2 ± 365.3 | 0.326 |
| IGFBP-3 (ng/mL)† | 17.1 [5.9]<br>18.5 ± 4.3 | 18.9 [3.4]<br>18.6 ± 2.4 | 0.687 |
| IGF-1/IGFBP-3† | 4.8 [3.2]<br>5.1 ± 2.2 | 4.4 [3.2]<br>4.9 [1.9] | 0.906 |

Data shown as median [interquartile range] and mean ± SD. P values obtained via Wilcoxon signed rank test.\*Data available in 15 women. † Data available in 17 women. IGF-1 and IGFBP represent morning fasting levels. GH levels were obtained every 10 minutes from 8 PM until 8 AM.

**Figure 5.**
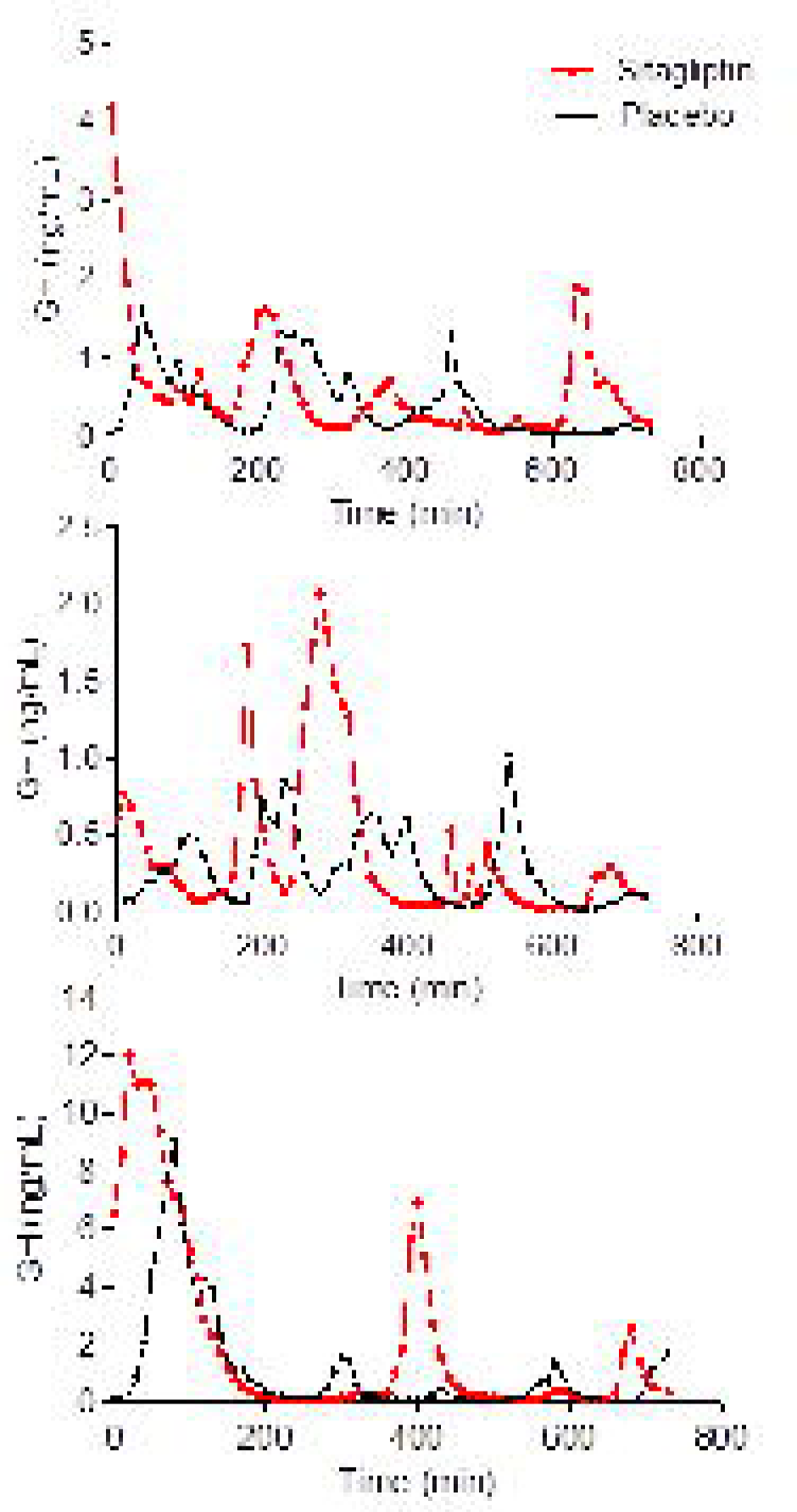
Representative plots of overnight GH levels from 3 subjects during placebo and sitagliptin treatment. GH samples are obtained every 10 minutes from 8 PM to 8 AM. The plots illustrate the high inter-individual variability but low intra-individual variability in overnight GH secretion.

### Effect of sitagliptin on vascular parameters and conduit artery vascular function

Sitagliptin increased heart rate during the 75-gram OGTT summarized as average over time (mean difference 2 bpm, 95% CI [0.01, 4.06]; p<0.05) and during the inpatient study day (from 67.4 ± 9.3 to 71.3 ± 6.9 bpm; p<0.05). One month of sitagliptin did not affect inflammatory or fibrinolytic markers. Sitagliptin also had no effect on conduit artery vascular function during reactive hyperemia. (**Table 4**) A limited number of women (N=3) met criteria for assessment of endothelium-independent vasodilation using nitroglycerin and agreed to its administration. Sitagliptin did not influence vasodilation after 0.4 mg sublingual nitroglycerin (36.5% ± 13.7 after placebo vs. 36.4% ± 9.0 after sitagliptin, p>0.05).

**Table 4.** Effect of sitagliptin on vascular parameters and conduit artery vascular function.

| Variable (N=17) | Placebo | Sitagliptin | P-Value |
| --- | --- | --- | --- |
| Systolic Blood Pressure (mm Hg) | 114.7 [18.7]<br>117.8 ± 11.6 | 115.0 [16.2]<br>115.6 ± 8.7 | 0.518 |
| Diastolic Blood Pressure (mmHg) | 75.0 [8.9]<br>73.8 ± 7.4 | 73.0 [11.0]<br>73.7 ± 7.8 | 1.000 |
| Heart rate (beats per minute) | 67.0 [14.2]<br>67.4 ± 9.3 | 68.0 [7.0]<br>71.3 ± 6.9 | **0.046** |
| Baseline artery diameter (mm) | 3.14 [0.55]<br>3.16 ± 0.43 | 3.12 [0.62]<br>3.13 ± 0.37 | 0.796 |
| Baseline flow integral (m) | 0.12 [0.04]<br>0.15 ± 0.08 | 0.17 [0.07]<br>0.18 ± 0.08 | 0.266 |
| Peak FMD (mm) | 3.53 [0.66]<br>3.58 ± 0.47 | 3.48 [0.54]<br>3.53 ± 0.36 | 0.344 |
| Time of peak FMD (sec) | 59.00 [37.82]<br>60.71 ± 30.80 | 64.66 [46.56]<br>75.55 ± 35.02 | 0.356 |
| Diameter increase (mm) | 0.39 [0.26]<br>0.43 ± 0.16 | 0.40 [0.23]<br>0.40 ± 0.16 | 0.705 |
| Flow-mediated vasodilation (%) | 12.42 [8.28]<br>13.63 ± 5.27 | 14.24 [8.45]<br>13.00 ± 5.34 | 0.943 |
| Peak VTI (m)* | 1.13 [0.34]<br>1.12 ± 0.23 | 1.14 [0.27]<br>1.13 ± 0.32 | 0.717 |
| VTI, fold increase* | 8.37 [3.49]<br>8.34 ± 2.66 | 7.26 [4.35]<br>7.38 ± 2.79 | 0.234 |
| tPA activity (IU/mL) | 0.21 [0.40]<br>0.40 ± 0.47 | 0.23 [0.82]<br>0.44 ± 0.46 | 0.831 |
| PAI-1 antigen (ng/mL)* | 8.78 [7.33]<br>10.96 ± 7.51 | 9.72 [4.77]<br>11.27 ± 8.05 | 0.959 |
| hsCRP (mg/L) | 3.51 [4.40]<br>4.43 ± 4.00 | 2.90 [3.45]<br>4.92 ± 5.36 | 0.619 |
| F2 Isoprostanes (pg/mL)* | 55.74 [44.02]<br>68.16 ± 45.69 | 50.02 [42.26]<br>62.01 ± 30.25 | 0.836 |

Data shown as median [interquartile range] and mean ± SD. P values obtained via Wilcoxon signed rank test. \*Data available in 16 women.

### Effect of sitagliptin on body composition and characteristics of PCOS

One month of sitagliptin therapy decreased VAT mass (p=0.02 vs. placebo) and volume (p=0.02 vs. placebo), but did not significantly affect weight or overall percent fat (**Table 5**). Sitagliptin decreased total cholesterol (from 168.8 ± 26.3 to 162.5 ± 22.2 mg/dL; p=0.02) largely due to a decrease in LDL (from 101.9 ± 23.8 to 96.2 ±21.5 mg/dL; p=0.06). One-month treatment with sitagliptin did not influence estradiol or testosterone levels (**Table 5**). Sitagliptin did not affect health-related quality of life domains (emotions, body hair, weight, infertility and menstrual problems), as assessed by PCOS Questionnaire administered on the last day of each therapy (p=1.00 vs. placebo for each domain).

**Table 5.** Effect of sitagliptin on body composition and characteristics of PCOS.

| Variable (N=17) | Placebo | Sitagliptin | P-value |
| --- | --- | --- | --- |
| Weight (kg) | 81.0 [28.0]<br>90.4 ± 21.7 | 81.6 [27.1]<br>90.2 ± 22.1 | 0.619 |
| Body Mass Index (kg/m<sup>2</sup>) | 32.1 [10.7]<br>34.3 ± 6.4 | 32.4 [10.4]<br>34.2 ± 6.4 | 0.586 |
| Waist Circumference (cm) | 105.9 [27.7]<br>105.5 ± 18.0 | 105.0 [18.7]<br>104.0 ± 15.4 | 0.381 |
| Waist-Hip Ratio* | 0.89 [0.12]<br>0.88 ± 0.07 | 0.87 [0.10]<br>0.87 ± 0.07 | 0.326 |
| Percent Body Fat (%) | 46.2 [5.2]<br>46.8 ± 5.4 | 45.8 [6.5]<br>46.3 ± 5.0 | 0.058 |
| Fat (kg) | 36.4 [16.9]<br>41.6 ± 14.3 | 35.5 [18.0]<br>40.9 ± 14.5 | 0.093 |
| Lean Body Mass (kg) | 43.9 [10.1]<br>45.6 ± 7.4 | 44.6 [10.4]<br>45.8 ± 7.9 | 0.758 |
| VAT Volume (cm<sup>3</sup>) | 1199.0 [744.0]<br>1210.6 ± 743.0 | 959.0 [820.5]<br>1118.4 ± 752.5 | **0.022** |
| VAT Mass (g) | 1131.0 [702.0]<br>1141.9 ± 700.7 | 904.0 [774.0]<br>1055.1 ± 710.1 | **0.022** |
| Android (% fat)† | 54.1 [9.2]<br>54.0 ± 7.6 | 51.9 [11.2]<br>52.8 ± 7.0 | **0.037** |
| Bioavailable Testosterone (ng/dL)* | 23.2 [11.5]<br>22.9 ± 10.3 | 16.1 [12.0]<br>19.8 ± 10.5 | 0.109 |
| Total Testosterone (ng/dL) | 43.0 [25.0]<br>48.4 ± 24.3 | 36.0 [31.0]<br>45.1 ± 27.5 | 0.266 |
| SHBG (nmol/L)* | 25.5 [20.0]<br>33.4 ± 21.1 | 31.5 [18.8]<br>30.7 ± 15.7 | 0.325 |
| Estradiol (pg/mL) | 212.9 [140.6]<br>233.5 ± 118.9 | 210.2 [91.9]<br>214.6 ± 71.4 | 0.407 |
| Total Cholesterol (mg/dL) | 169.0 [52.5]<br>168.8 ± 26.3 | 165.0 [32.0]<br>162.5 ± 22.2 | **0.020** |
| LDL (mg/dL) | 93.0 [45.0]<br>101.9 ± 23.8 | 93.0 [37.5]<br>96.2 ± 21.5 | 0.055 |
| HDL (mg/dL) | 49.0 [18.5]<br>53.8 ± 18.3 | 51.0 [15.0]<br>52.8 ± 15.3 | 0.462 |
| Triglycerides (mg/dL) | 65.0 [39.0]<br>64.8 ± 24.0 | 53.0 [58.5]<br>67.6 ± 36.9 | 0.887 |

Data shown as median [interquartile range] and mean ± SD. P values obtained via Wilcoxon signed rank test. \*Data available in 16 women. † Android (% fat) represents the %total fat within the android (central) region, as opposed to the gynoid (hip and thigh) region.

### Association between DPP4 inhibition and GH levels, VAT, and HOMA-IR

Sitagliptin did not increase overnight GH secretion or lower insulin resistance. Greater DPP4 inhibition in response to sitagliptin, however, correlated with GH levels, VAT and HOMA-IR. The degree of DPP4 inhibition with sitagliptin correlated with GH secretion, such that DPP4 inhibition was associated with overnight GH secretion (GH AUC: r_s_= 0.60, p=0.02; mean GH: r_s_= 0.59, p=0.02). DPP4 inhibition after sitagliptin was also associated with VAT mass (r_s_= −0.66; p<0.01), and HOMA-IR (r_s_=-0.64; p=0.01). The correlations between DPP4 inhibition and GH AUC, VAT, and HOMA-IR were no longer significant after controlling for BMI (GH AUC r_s_=0.25, p=0.38; VAT r_s_=-0.31, p=0.25; HOMA-IR r_s_=-0.15; p=0.61). Women with higher DPP4 inhibition during sitagliptin treatment, i.e. those in whom the percent DPP4 inhibition (defined as [1-(DPP4 activity during sitagliptin/DPP4 activity during placebo)]*100) was above the median for the women analyzed, had higher overnight GH secretion and lower VAT mass and HOMA-IR. **Figure 6** compares values between those with higher DPP4 inhibition and lower DPP4 inhibition within the sitagliptin treatment. Similarly, women with greater DPP4 inhibition also had lower BMI (30.3 ± 3.4 vs. 38.5 ± 6.3 kg/m^2^; p<0.01).

**Figure 6.**
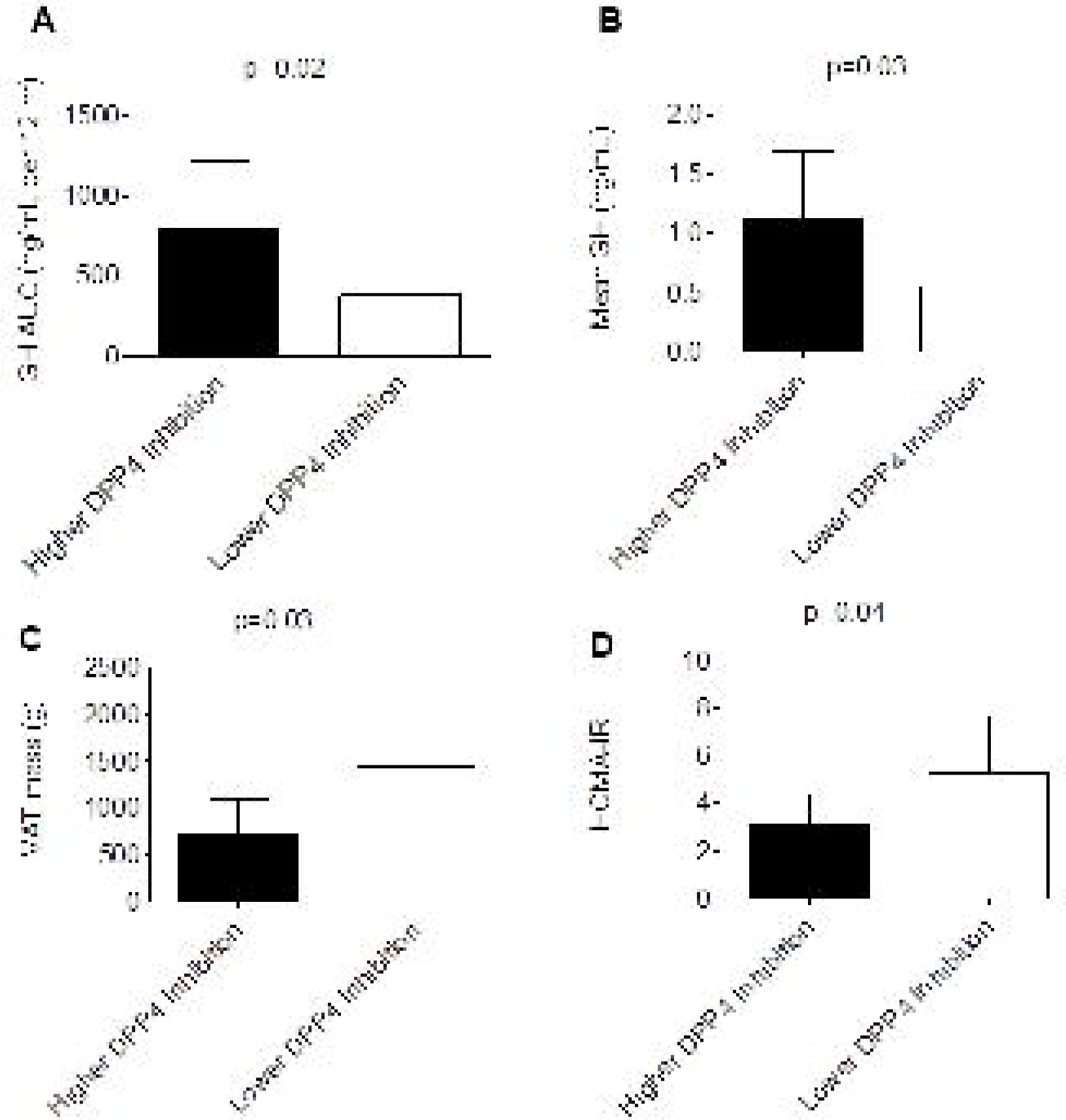
Women with a higher percent DPP4 inhibition had higher overnight GH secretion (A and B, N=16) and lower VAT mass (C, N=17) and HOMA-IR (D, N=17) after one-month treatment with sitagliptin. Data were obtained on the inpatient visit day and are expressed as mean ± SD. P values were calculated using the Wilcoxon rank sum test. Higher percent DPP4 inhibition is defined as percent DPP4 inhibition at or greater than the median for the 17 women included in the analysis, where percent DPP4 inhibition = [1-(DPP4 activity during sitagliptin/DPP4 activity during placebo)]*100. Comparisons between higher and lower DPP4 inhibition were not significant after controlling for BMI (GH AUC p=0.27; mean GH p=0.24; VAT p=0.67; HOMA-IR p=0.66)

### Safety

One woman was hospitalized with gallstone-induced pancreatitis after two doses of sitagliptin. One woman reported intermittent abdominal pain while taking sitagliptin and another reported dizziness. One woman taking sitagliptin reported dizziness towards the end of the OGTT, another woman taking sitagliptin developed headache during the OGTT. There were no reported instances of hypoglycemia while taking sitagliptin. Three woman reported nausea, decreased appetite, and dizziness while taking placebo. One woman developed nausea during the OGTT while taking placebo, and one experienced emesis. One woman developed shingles while taking placebo. One woman developed headache, nausea, and dizziness following nitroglycerin.

## Discussion

This study tested the hypothesis that one-month treatment with the DPP4 inhibitor sitagliptin would potentiate overnight GH secretion in overweight women with PCOS and improve glucose levels. As expected, sitagliptin lowered blood glucose by increasing GLP-1 levels and enhancing early insulin secretion following ingestion of oral glucose. Sitagliptin did not increase mean overnight GH but did enhance GH half-life. Body composition also improved during sitagliptin, as evidenced by a decrease in VAT, while weight and BMI remained stable. These findings indicate that DPP4 inhibition with sitagliptin decreases the maximal glucose response to OGTT, improves body composition, and increases GH half-life in overweight women with PCOS.

Altered GLP-1 and insulin secretion following oral glucose ingestion has previously been reported even in lean, glucose-tolerant women with PCOS. Women with PCOS have higher levels of c-peptide and tend to have higher levels of insulin secretion particularly in the first hour after glucose ingestion. While active GLP-1 levels are similar in women with PCOS and healthy volunteers early on, they are lower in women with PCOS after one hour.(34) The glucagon response to hypoglycemia is also potentiated in obese women with PCOS as compared to matched controls.(35) We hypothesized that sitagliptin, with its ability to potentiate post-prandial GLP-1 secretion and suppress glucagon secretion, would mitigate these undesirable post-glucose changes. We observed enhanced GLP-1 secretion during sitagliptin, but no changes in glucagon levels.

Only one previous study has evaluated the effect of sitagliptin on glucose homeostasis in women with PCOS. Ferjan et al. conducted a 12-week study of obese women with PCOS in which women were randomized to open label sitagliptin 100 mg daily with lifestyle intervention or lifestyle intervention alone following metformin withdrawal.(36) The investigators found that sitagliptin improved beta cell function and even prevented the conversion from impaired glucose tolerance to type 2 diabetes mellitus in three of the women. The investigators did not appreciate a significant decrease in glucose levels during OGTT. In contrast, we observed that sitagliptin enhanced GLP-1 secretion following oral glucose ingestion which resulted in enhanced early insulin secretion and a significant decrease in the blood glucose rise following oral glucose ingestion. Our relatively more homogeneous study population and cross-over design likely enabled us to detect better an improvement in blood glucose levels.

The effect of sitagliptin on post-prandial GH secretion has not previously been investigated. It is well known that oral glucose ingestion quickly inhibits GH secretion before a late stimulatory effect on GHRH-mediated GH levels. Relatively little is known, however, about the metabolic factors that influence GH secretion following oral glucose. Grottoli et al. demonstrated that early GH inhibition following glucose ingestion is lost in obesity but the late stimulatory effect is maintained.(37) Iranmanesh et al further demonstrated that nadir and peak rebound GH secretion is influenced by VAT and SHBG in obese men.(38) Potential mechanisms by which sitagliptin might alter the GH secretion in response to an OGTT in women with PCOS include enhanced late GH secretion due to decreased degradation of the GHRH stimulus or mitigation of the known inhibitory effects of hyperglycemia and free fatty acids on GH secretion. Although sitagliptin lowered blood glucoses following oral glucose ingestion, we did not observe any effect of sitagliptin on free fatty acids or on GH secretion. Skov et al also demonstrated that intravenous GLP-1 infusion in men reduces IGFBP-1 and tended to increase IGF-1 bioactivity.(39) We did see an increase in GLP-1 and early insulin secretion, but this did not significantly influence IGFBP-1 levels or free IGF-1 levels. Thus, it is unlikely that alterations in insulin secretion, blood glucoses, IGFBP-1, or GLP-1 levels contribute to the altered post-glucose GH secretion described in obesity. Furthermore, IGF-1 did not contribute to the observed changes in glucose metabolism in our study participants.

Our study investigates a novel potential strategy to enhance endogenous GH secretion in a population of patients demonstrated to have diminished GH levels. Obese individuals have diminished GH secretion, characterized by fewer GH pulses and shorter half-life duration.(40) De Boer et. al. observed impaired overnight GH secretion in PCOS patients, irrespective of weight and degree of insulin resistance.(12) We found that one month of sitagliptin therapy influences overnight GH secretion by increasing GH half-life. The inter-pulse interval, which represents the time interval between secretion events and is partially dependent upon half-life in addition to the previous inter-pulse interval and secretion event area, was also increased.(28,29) Sitagliptin did not increase overnight GH levels. Although overnight GH secretion correlated with DPP4 inhibition after sitagliptin. this was not the case after controlling for BMI.

Our study was not designed to determine the mechanism by which sitagliptin influences the overnight GH secretory profile. Possible mechanisms include decreased degradation of the GHRH stimulus by DPP4, or increased GH as an indirect result of decreased VAT. In support of the first mechanism, Makimura et. al. reported that twelve months of the DPP4-resistant GHRH analogue, tesamorelin, increases IGF-1 and selectively decreases VAT in obese subjects with reduced GH secretion.(41) In contrast, we did not find that sitagliptin affected IGF-1 levels, yet VAT was still selectively affected. The lack of effect of sitagliptin on IGF-1 levels is not surprising given that sitagliptin did not affect overnight GH secretion. In support of the second mechanism, Vahl et al found that abdominal obesity was the single most significant determinant of pulsatile GH release, as determined by twenty-four hour GH profile.(42) Pijl et. al. studied the effect of caloric restriction and 50% loss of body weight over a 3 to 5 month period in obese women and observed an increase in mean 24 hour GH secretion.(6) Similar to our findings, they observed an increase in GH half-life with no effect on IGF-1 levels. This report is consistent with our observation of a smaller yet significant decrease in VAT and increase in GH half-life after only one month of sitagliptin in our subjects. Dichtel LE et al reported that in overweight and obese women, increases in bioactive IGF-1, rather than total IGF-1, drive GH-mediated body composition changes in the short-term.(43) A decrease in GH binding protein may also have played a role in the observed changes in GH half-life and inter-pulse interval, as levels of GH binding protein directly correlate with VAT.(44) Lastly, another mechanism possibly contributing to the decrease in VAT may be increased sympathetic activity. We have previously reported that sitagliptin increases sympathetic activity during concurrent angiotensin converting-enzyme (ACE) inhibition by decreasing degradation of the DPP4 and ACE substrate substance P.(45) We did observe an increase in heart rate following sitagliptin during the OGTT.

Others have reported that the addition of sitagliptin to metformin therapy improves body composition in women with PCOS. Yan J et al reported that the addition of sitagliptin to metformin treatment reduces VAT in patients with type 2 diabetes and non-alcoholic fatty liver disease.(46) Ferjan et. al. similarly found that sitagliptin prevents weight regain, as compared to metformin alone, in a 12-week prospective open label study in 24 obese women with PCOS previously treated with liraglutide.(47) Thus, a growing body of evidence suggests that sitagliptin selectively reduces VAT. Despite the improvements in VAT and GH secretion, we did not observe an improvement in any of the clinical signs or symptoms experienced by women with PCOS, as reported in a validated PCOS questionnaire. Longer duration of therapy would likely be necessary to detect a change in these parameters. Similarly, androgen levels were not affected by sitagliptin. We would expect androgen levels to improve in association with an improvement in insulin resistance, and the latter was not observed.

Our study further evaluates cardiovascular endpoints, such as vascular function and fibrinolysis, which may be affected by enhanced GH secretion. GH directly improves vascular function through several mechanisms, including activation of the GH receptor and downstream IGF-1 secretion (48), as well as GH receptor- and IGF-1-independent effects.(49) Li et. al. reported that supra-physiologic circulating concentrations GH of approximately 33 ng/mL were necessary to increase forearm blood flow.(1) Similarly, Napoli et al reported that intra-arterial GH infusion, which raised local GH to approximately 40 ng/mL, increased forearm blood flow.(2) We previously reported that sitagliptin increases forearm vasodilation, measured by strain gauge plethysmography, in healthy women as assessed during stimulated GH secretion with peak levels averaging 11.6 mg/dL.(11) In the present study, we did not observe any effect of sitagliptin on conduit artery vascular function, however. Mean overnight GH levels in this study in patients with PCOS were significantly lower than in our prior study in healthy women. The changes in GH secretion induced by sitagliptin may have been insufficient to detect an effect on forearm vasodilation. Other potential factors, such as increased sympathetic activity due to decreased degradation of other DPP4 substrates such as substance P and neuropeptide Y, may also have masked any GH-induced changes in vascular function.(45,50) It is also notable that we did not observe any increase in vasodilation despite elevated GLP-1 levels during sitagliptin. This is consistent with our previous observation that GLP-1 infusion into the forearm vasculature does not increase vasodilation.(51)

We report for the first time the effect of chronic sitagliptin treatment on tPA activity and PAI-1 antigen levels. Despite the observed decrease in VAT and cholesterol, there was no significant effect of sitagliptin on PAI-1. This contrasts with the data of Tani et al who found that eight-week treatment with vildagliptin decreased triglycerides and PAI-1 in poorly controlled type 2 diabetic patients.(52) The investigators suggested that improved lipid metabolism resulted in down-regulation of PAI-1 production from the vascular endothelial cells. Triglyceride and PAI-1 levels in the women in the current study were one-third of the levels in their study population, thus making it difficult to detect further down-regulation of PAI-1 production. We have previously demonstrated that GH affects tPA activity levels through the GH receptor and downstream IGF-1 secretion,(11) but we did not observe any change in IGF-1 or tPA activity in this study. We also did not see any effect of sitagliptin on inflammation, whereas Makdissi A et al observed an anti-inflammatory effect of 100 mg daily sitagliptin on CRP, IL-6 and free fatty acids after 12 weeks in patients with type 2 diabetes.(53) Differences in study populations or treatment duration could account for these different observations.

Our study has limitations. PCOS is a heterogeneous disorder; women may be lean or overweight and may or may not have insulin resistance. We studied a relatively homogenous subset of women with PCOS and our results may not be generalizable to all women with PCOS. We chose to limit our study population to those women with a BMI of at least 25 kg/m^2^ and to exclude participants who were dieting with weight loss or titrating metformin therapy. This subset of women with PCOS tends to represent the subset who are prescribed metformin in order to improve glucose metabolism. We were interested in how sitagliptin influenced glucose metabolism in overweight women with PCOS and we were limited to use of a manual glucometer for glucose measurements, though the same glucometer was used for each woman across study days. We also did not include baseline endogenous GH secretory capacity as a selection criterion for the women studied, as is done in many clinical trials evaluating the effect of an intervention on GH secretion. If we had pre-selected for a relatively GH-deficient population, we may have seen a more prominent effect of sitagliptin on overnight GH secretion. We were unable to standardize timing of the endpoint measurements to womens’ menstrual cycles, as most women had infrequent menstrual cycles. This is particularly relevant in our assessments of overnight GH secretion and vascular function. We did not observe a difference in estradiol levels across study treatments, however. Our treatment duration was one month, which may not have been sufficient to appreciate any effect of sitagliptin on clinical symptoms, or inflammatory or fibrinolytic markers. We chose to use the current FDA-approved daily dose of sitagliptin, however we have previously demonstrated that 100 mg daily of sitagliptin is not as effective as single-dose 200 mg sitagliptin in inhibiting DPP4 activity in healthy individuals, and that sitagliptin 100 mg daily is less effective in inhibiting DPP4 activity in individuals with type 2 diabetes mellitus and hypertension than in healthy individuals.(54) A higher dose of sitagliptin resulting in maximal DPP4 inhibition may have yielded a greater treatment effect on mean GH and eliminated any effect of weight on response to sitagliptin. Our limited sample size may have limited our conclusions on overnight GH secretion parameters. We observed low intra-individual variability in the pattern of overnight GH secretion, however, and the cross-over design of the study would have increased our ability to detect treatment effects. Lastly, our study design did not enable us to determine the mechanism by which sitagliptin may enhance GH half-life.

In this study, we evaluated the previously unexplored effects of sitagliptin in overweight women with PCOS. We demonstrate for the first time that sitagliptin enhances GLP-1 levels and early insulin secretion and thereby decreases peak blood glucose following oral glucose ingestion in these women. Second, we demonstrate that sitagliptin improves body composition, through an undefined mechanism. As weight loss remains the cornerstone of treatment for PCOS, this is also a highly desirable effect. Lastly, this study is the first to suggest an off-target effect of sitagliptin on GH secretion in women with PCOS, potentiating GH half-life and the inter-pulse interval. This is relevant as endogenous GH secretion is diminished in women with PCOS. (12-14) These data are the first to support the use of oral DPP4 inhibitor therapy to address elevated blood glucoses and body composition in women with PCOS, and to probe the contribution of an altered somatotropic axis in the pathophysiology of this common disorder.

## Data Availability

Data are available upon request.

## Acknowledgements

We thank Anthony Dematteo, Zuofei Wang, and Aaron Falck for their technical assistance. We thank Drs. Kevin Niswender, Italo Biaggioni, and Melissa Wellons for their expertise and service on the Data and Safety Monitoring Committee.

## Notes

**Sources of Funding**: This research was supported by Vanderbilt Clinical and Translational Science Awards (CTSA) grant UL1 TR000445-06 from the NIH National Center for Advancing Translational Sciences. The Vanderbilt Hormone Assay and Analytical Services Core is supported by NIH grants DK059637 and DK020593. JKD was supported by K23HL11962 NHLBI/NIH and NJB by R01HL125426 from NHLBI/NIH.

### Competing Interest Statement

The authors have declared no competing interest.

### Clinical Trial

NCT02122380

### Funding Statement

This research was supported by Vanderbilt Clinical and Translational Science Awards (CTSA) grant UL1 TR000445-06 from the NIH National Center for Advancing Translational Sciences. The Vanderbilt Hormone Assay and Analytical Services Core is supported by NIH grants DK059637 and DK020593. JKD was supported by K23HL11962 NHLBI/NIH and NJB by R01HL125426 from NHLBI/NIH.

### Author Declarations

All relevant ethical guidelines have been followed and any necessary IRB and/or ethics committee approvals have been obtained.

Any clinical trials involved have been registered with an ICMJE-approved registry such as ClinicalTrials.gov and the trial ID is included in the manuscript.

